# The clinical spectrum of encephalitis in COVID-19 disease: the ENCOVID multicentre study

**DOI:** 10.1101/2020.06.19.20133991

**Authors:** Andrea Pilotto, Stefano Masciocchi, Irene Volonghi, Elisabetta del Zotto, Eugenio Magni, Valeria De Giuli, Francesca Caprioli, Nicola Rifino, Maria Sessa, Michele Gennuso, Maria Sofia Cotelli, Marinella Turla, Ubaldo Balducci, Sara Mariotto, Sergio Ferrari, Alfonso Ciccone, Fabrizio Fiacco, Alberto Imarisio, Barbara Risi, Alberto Benussi, Enrico Premi, Emanuele Focà, Francesca Caccuri, Matilde Leonardi, Roberto Gasparotti, Francesco Castelli, Gianluigi Zanusso, Alessandro Pezzini, Alessandro Padovani, on behalf of ENCOVID Study Group#

## Abstract

**Importance:** Several preclinical and clinical investigations have argued for nervous system involvement in SARS-CoV-2 infection. Some sparse case reports have described various forms of encephalitis in COVID-19 disease, but very few data have focused on clinical presentations, clinical course, response to treatment and outcomes yet.

**Objective:** to describe the clinical phenotype, laboratory and neuroimaging findings of encephalitis associated with SARS-CoV-2 infection, their relationship with respiratory function and inflammatory parameters and their clinical course and response to treatment.

**Design:** The ENCOVID multicentre study was carried out in 13 centres in northern Italy between February 20^th^ and May 31^st^, 2020. Only patients with altered mental status and at least two supportive criteria for encephalitis with full infectious screening, CSF, EEG, MRI data and a confirmed diagnosis of SARS-CoV-2 infection were included. Clinical presentation and laboratory markers, severity of COVID-19 disease, response to treatment and outcomes were recorded.

**Results:** Out of 45 cases screened, twenty-five cases of encephalitis positive for SARS-CoV-2 infection with full available data were included. The most common symptoms at onset were delirium (68%), aphasia/dysarthria (24%) and seizures (24%). CSF showed hyperproteinorrachia and/or pleocytosis in 68% of cases whereas SARS-CoV-2 RNA by RT-PCR resulted negative. Based on MRI, cases were classified as ADEM (n=3), limbic encephalitis (LE, n=2), encephalitis with normal imaging (n=13) and encephalitis with MRI alterations (n=7). ADEM and LE cases showed a delayed onset compared to the other encephalitis (p=0.001) and were associated with previous more severe COVID-19 respiratory involvement. Patients with MRI alterations exhibited worse response to treatment and final outcomes compared to other encephalitis.

**Conclusions and relevance:** We found a wide clinical spectrum of encephalitis associated with COVID19 infection, underlying different pathophysiological mechanisms. Response to treatment and final outcome strongly depended on specific CNS-manifestations.

**Question:** what are the phenotypes of encephalitis associated to SARS-CoV-2 infection?

**Findings:** 25 cases of encephalitis in SARS-CoV-2 infection were included in a prospective observational multi-centre study. Encephalitis cases in COVID-19 exhibited a wide heterogeneity in terms of clinical features, CSF, MRI findings, response to treatment and outcomes with 13 cases with normal MRI, 7 with heterogeneous MRI alterations and rarer ADEM/limbic encephalitis cases.

**Meaning:** heterogeneity of presentation, response to treatment and outcomes of encephalitis of COVID-19 underlines different pathophysiological mechanisms

## Introduction

SARS-CoV-2 infection is mainly characterized by respiratory tract symptoms with adverse outcomes mostly related to the severity of acute respiratory distress syndrome (ARDS). With the increasing number of confirmed cases and the accumulating clinical data, it is now well established that, in addition to the predominant respiratory symptoms, a significant proportion of COVID-19 patients experience neurological symptoms, including olfactory dysfunction,^1^ headache and confusion.^2,3^ SARS-CoV-2 infection has also the potential for specifically targeting the central nervous system (CNS) and some cases of encephalitis have been described in patients admitted to intensive care unit for severe respiratory distress.^4,5^

Few case reports also described the occurrence of encephalitis during SARS-CoV-2 infection independently from ARDS, arguing for a CNS inflammatory-mediated involvement.^6,7^ However, the neurotropism of SARS-CoV-2 infection and the mechanisms leading to specific brain involvement are still theme of debate.^8,9,10^ In fact, literature findings have been limited to few single-centre series while a full work-up for alternative causes of brain infections was often not performed.^11^

In order to investigate the relationship between COVID-19 and CNS involvement, a network was established involving 13 neurological units in Lombardy and Veneto, covering an area with the highest number of COVID-19 cases observed in Italy so far.^12^

The aim of the study was to identify all cases of encephalitis associated with SARS-CoV-2 infection, to analyse their relationship with respiratory function and inflammatory parameters and to describe their clinical, laboratory and neuroimaging findings, the clinical course and response to treatment.

## METHODS

### Patients

Data were collected in the setting of a multicentre, ongoing, hospital-based, longitudinal cohort study on encephalitis during the COVID-19 pandemic (ENCOVID-study). All patients consecutively admitted to the participating hospitals between February 20^th^ and May 31^st^ 2020, with symptoms suggestive of encephalitis were screened for inclusion. The case definition included any person aged ≥18 admitted to hospital with altered mental status (defined as decreased or altered level of consciousness, lethargy or personality changes) lasting ≥24 hours and the presence of two or more of the following criteria^13^: i) generalized or partial seizures not fully attributable to a pre-existing epilepsy ii) new onset of focal neurologic findings iii) CSF white blood cell count ≥5/cubic mm^3^ iv) abnormality of brain parenchyma on neuroimaging suggestive of encephalitis that was either new from prior studies or appears acute in onset v) abnormality on electroencephalography consistent with encephalitis and not attributable to another cause.

Fever was not considered as supportive feature for encephalitis (as indicated by standard criteria^13^), as it is highly prevalent in COVID-19 disease.^14^ Demographic information, clinical manifestations, laboratory findings, EEG, neuroimaging, treatment and outcomes were extracted from medical records using a standardized anonymized data collection form by study physicians and checked by senior investigators at each centre.

The Institutional Ethical Standards Committee on human experimentation at Brescia University Hospital provided approval for the study (NP 4067, approved May 8^th^, 2020).

### COVID-19 diagnosis and stratification

Laboratory confirmation of SARS-CoV-2 infection was carried out by RT PCR procedure on throat-swab and nasopharyngeal specimens in all patients. In case of high clinical suspicion of SARS-CoV-2 infection and negative test results on nasopharyngeal and oropharyngeal swabs, testing of lower respiratory samples (bronchoalveolar lavage fluid obtained by bronchoscopy) was performed. In all patients, routine blood examinations included complete blood count, coagulation profile, serum biochemical tests, including renal and liver function and lactate dehydrogenase. The illness severity of COVID-19 was defined according to the Brescia COVID Respiratory Severity Scale (BCRSS) for COVID-19^15^ and according to the quick Sequential Organ Failure Assessment (qSOFA).^14^ Respiratory function at admission, during hospitalization and at discharge was defined by no support of oxygen therapy, low-flux oxygenation (FiO2 ≤50%) and high-flux oxygenation (FiO2 ≥50%).

### Encephalitis assessment

First-line testing (supplementary table) included all commonly recognized causes of encephalitis according to current guidelines.^13,16^

Brain MRI, standard EEG and CSF analyses were performed in all patients if no contraindicated from general medical conditions. Only patients with full data available for MRI, EEG and CSF were included in the final analyses. When possible, encephalitis was classified as acute demyelinating encephalomyelitis (ADEM)/acute necrotizing encephalitis (ANE)^16,17,18^ or limbic encephalitis (LE)^16^ based on clinical and instrumental data.^13^

### Treatment and outcome measures

Treatment of COVID-19 infection was decided by an infectious disease specialist at each centre and included chloroquine/hydroxycloroquine or antiviral treatment, such as lopinavir/ritonavir 200/50 mg 2 BID, darunavir 800 mg 1 QD + ritonavir 100 mg QD, or darunavir/cobicistat 800/150 mg QD. Treatment of respiratory insufficiency was managed according to standardized protocols for high-flow oxygenation or intensive care unit admission. Specific treatments for encephalitis included i) antibiotic treatment for suspected meningitis ii) antiviral treatment in case of suspected viral encephalitis iii) high-dose steroid treatment and iv) immunoglobulins/plasmapheresis.

Modified Rankin Scale (mRS)^19^ was administered both at admission and at discharge. Poor outcome was defined as death or severe disability (mRS>4). A “positive response” to specific treatment was defined as an improvement of at least two points on the mRS scale.^19^

### Statistical analyses

Data are presented as mean ± standard deviation for continuous variables and number (%) for categorical variables. For subgroup comparisons, we used the Fisher exact test and ANOVA test, when appropriate.

## RESULTS

The study network recruited 45 cases of encephalitis starting from February 20^th^ to May 31^st^ 2020. Of these, 32 cases were affected by SARS-CoV-2 virus infection. The remaining 13 SARS-CoV-2 negative patients (7 males and 6 females, mean age 53.2 ± 24 years) were characterised by 3 limbic encephalitis (2 with positive CSF LG1 antibodies), 2 herpes virus encephalitis, one ADEM, one tuberculosis-related meningo-encephalitis and 6 encephalitis of unknown cause.

Six cases of encephalitis in SARS-CoV-2 infection were excluded because lacking of complete clinical description, EEG, MRI or CSF analyses (Supplementary Figure 1). One case of HSV-1 severe encephalitis in COVID-19 was excluded because of early death and no MRI assessment. Twenty-five cases of encephalitis (15 males and 10 females, mean age 65.9 ± 9.6, range 50-84 years) were included in the final analyses (Supplementary Figure 1). The number of patients argues for an estimated incidence of 58/100.000 cases, considering the total of 43,139 COVID-19 cases identified within the same provinces from February 20^th^ and May 31^st^ 2020.

At admission, fifteen patients (60%) presented with COVID-19 related symptoms including fever, cough, dyspnoea and fatigue. At chest radiography, 24 cases showed interstitial pneumonia characterized by different degree of clinical severity as measured by the BCRSS (1.6 ± 1.3) and by qSOFA (1.2 ± 0.8). Ten (40%) cases required low-flux O2 and five high-flux oxygenation. During hospitalisation, five patients required the use of non-invasive ventilation, and four patients required invasive ventilation.

Hydroxychloroquine was administrated to 19 (76%) of cases while antiviral treatment was administered to 11 cases. Laboratory findings on admission showed anaemia in 14 patients, leucocytosis in 3, lymphocytopenia in 5, increased C-reactive protein (CRP) in 15, D-Dimer in 18 and fibrinogen in 13 (Table 1).

**Table 1.**
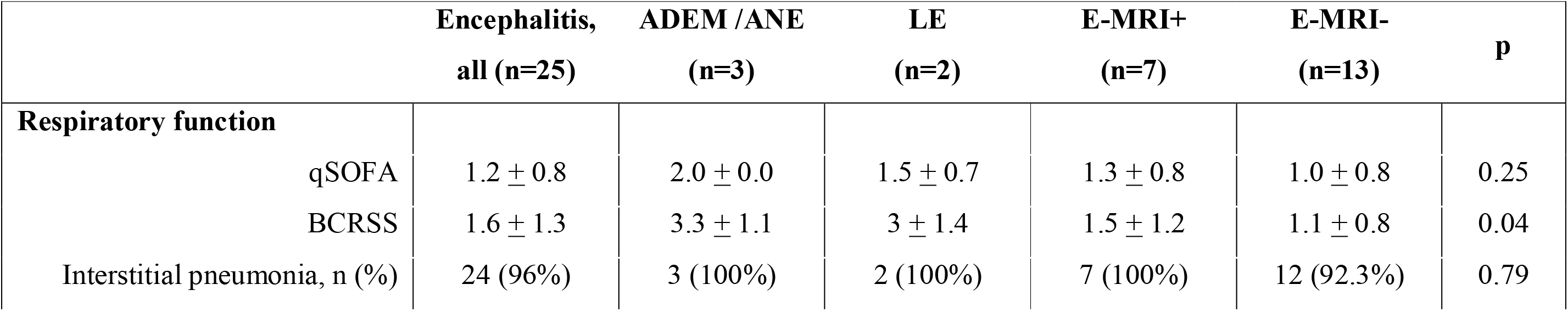
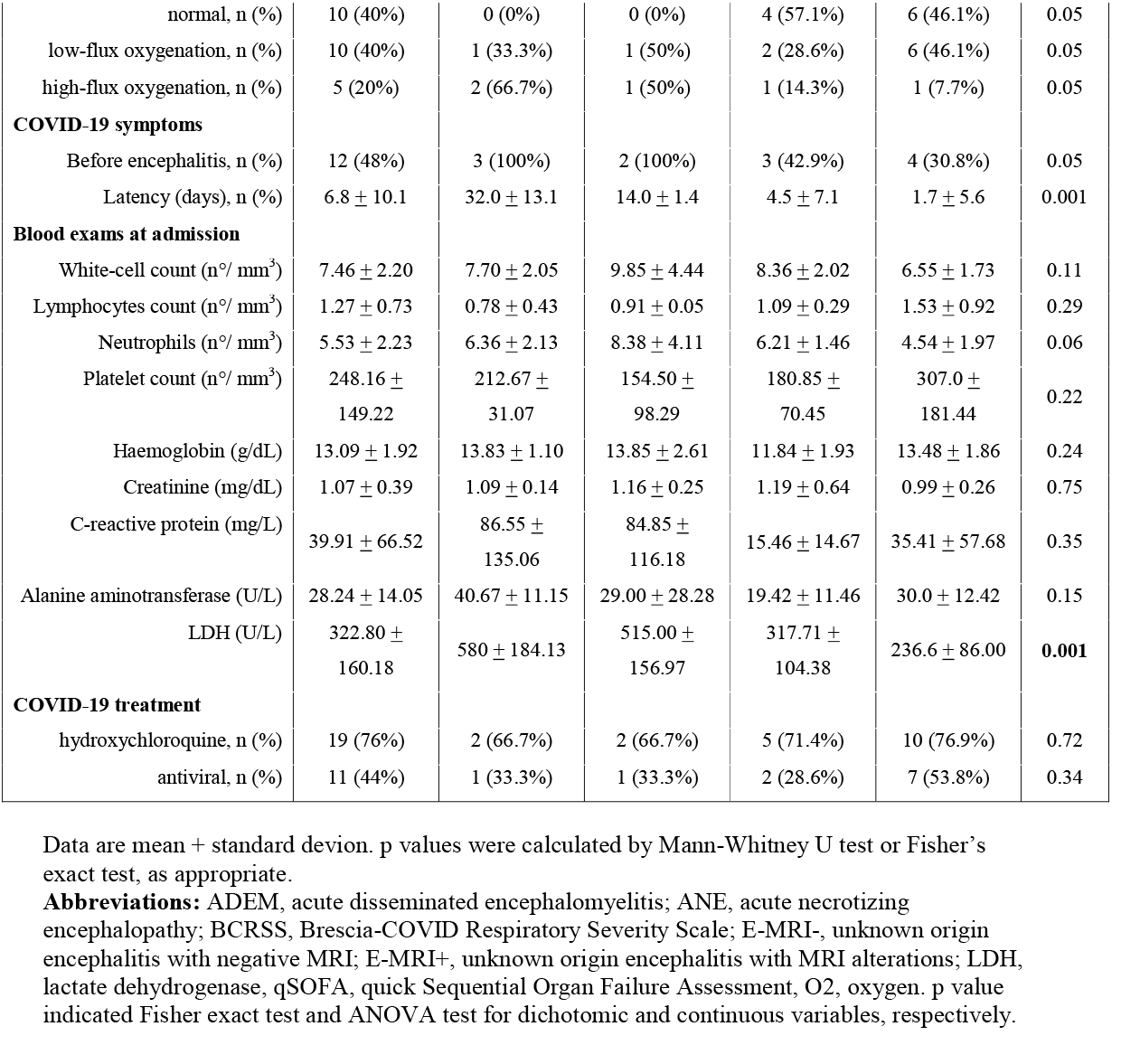
Respiratory function and SARS-CoV-2 infection characteristic of patients with encephalitis at admission.

|  | Encephalitis,<br>all (n=25) | ADEM /ANE<br>(n=3) | LE<br>(n=2) | E-MRI+<br>(n=7) | E-MRI-<br>(n=13) | p |
| --- | --- | --- | --- | --- | --- | --- |
| **Respiratory function** |  |  |  |  |  |  |
| qSOFA | 1.2 ± 0.8 | 2.0 ± 0.0 | 1.5 ± 0.7 | 1.3 ± 0.8 | 1.0 ± 0.8 | 0.25 |
| BCRSS | 1.6 ± 1.3 | 3.3 ± 1.1 | 3 ± 1.4 | 1.5 ± 1.2 | 1.1 ± 0.8 | 0.04 |
| Interstitial pneumonia, n (%) | 24 (96%) | 3 (100%) | 2 (100%) | 7 (100%) | 12 (92.3%) | 0.79 |
| normal, n (%) | 10 (40%) | 0 (0%) | 0 (0%) | 4 (57.1%) | 6 (46.1%) | 0.05 |
| low-flux oxygenation, n (%) | 10 (40%) | 1 (33.3%) | 1 (50%) | 2 (28.6%) | 6 (46.1%) | 0.05 |
| high-flux oxygenation, n (%) | 5 (20%) | 2 (66.7%) | 1 (50%) | 1 (14.3%) | 1 (7.7%) | 0.05 |
| **COVID-19 symptoms** |  |  |  |  |  |  |
| Before encephalitis, n (%) | 12 (48%) | 3 (100%) | 2 (100%) | 3 (42.9%) | 4 (30.8%) | 0.05 |
| Latency (days), n (%) | 6.8 ± 10.1 | 32.0 ± 13.1 | 14.0 ± 1.4 | 4.5 ± 7.1 | 1.7 ± 5.6 | 0.001 |
| **Blood exams at admission** |  |  |  |  |  |  |
| White-cell count (n°/ mm<sup>3</sup>) | 7.46 ± 2.20 | 7.70 ± 2.05 | 9.85 ± 4.44 | 8.36 ± 2.02 | 6.55 ± 1.73 | 0.11 |
| Lymphocytes count (n°/ mm<sup>3</sup>) | 1.27 ± 0.73 | 0.78 ± 0.43 | 0.91 ± 0.05 | 1.09 ± 0.29 | 1.53 ± 0.92 | 0.29 |
| Neutrophils (n°/ mm<sup>3</sup>) | 5.53 ± 2.23 | 6.36 ± 2.13 | 8.38 ± 4.11 | 6.21 ± 1.46 | 4.54 ± 1.97 | 0.06 |
| Platelet count (n°/ mm<sup>3</sup>) | 248.16 ±<br>149.22 | 212.67 ±<br>31.07 | 154.50 ±<br>98.29 | 180.85 ±<br>70.45 | 307.0 ±<br>181.44 | 0.22 |
| Haemoglobin (g/dL) | 13.09 ± 1.92 | 13.83 ± 1.10 | 13.85 ± 2.61 | 11.84 ± 1.93 | 13.48 ± 1.86 | 0.24 |
| Creatinine (mg/dL) | 1.07 ± 0.39 | 1.09 ± 0.14 | 1.16 ± 0.25 | 1.19 ± 0.64 | 0.99 ± 0.26 | 0.75 |
| C-reactive protein (mg/L) | 39.91 ± 66.52 | 86.55 ±<br>135.06 | 84.85 ±<br>116.18 | 15.46 ± 14.67 | 35.41 ± 57.68 | 0.35 |
| Alanine aminotransferase (U/L) | 28.24 ± 14.05 | 40.67 ± 11.15 | 29.00 ± 28.28 | 19.42 ± 11.46 | 30.0 ± 12.42 | 0.15 |
| LDH (U/L) | 322.80 ±<br>160.18 | 580 ± 184.13 | 515.00 ±<br>156.97 | 317.71 ±<br>104.38 | 236.6 ± 86.00 | **0.001** |
| **COVID-19 treatment** |  |  |  |  |  |  |
| hydroxychloroquine, n (%) | 19 (76%) | 2 (66.7%) | 2 (66.7%) | 5 (71.4%) | 10 (76.9%) | 0.72 |
| antiviral, n (%) | 11 (44%) | 1 (33.3%) | 1 (33.3%) | 2 (28.6%) | 7 (53.8%) | 0.34 |

Data are mean + standard deviation. p values were calculated by Mann-Whitney U test or Fisher's exact test, as appropriate.
**Abbreviations:** ADEM, acute disseminated encephalomyelitis; ANE, acute necrotizing encephalopathy; BCRSS, Brescia-COVID Respiratory Severity Scale; E-MRI-, unknown origin encephalitis with negative MRI; E-MRI+, unknown origin encephalitis with MRI alterations; LDH, lactate dehydrogenase, qSOFA, quick Sequential Organ Failure Assessment, O2, oxygen. p value indicated Fisher exact test and ANOVA test for dichotomic and continuous variables, respectively.

Onset of neurological symptoms was concomitant with COVID-19 symptomatology in 11 (44%) cases while in 12 cases (48%) neurological onset followed COVID-19 symptoms with a median of 8 (IQR 5 −12) days. In 2 cases neurological symptoms preceded the onset of COVID-19 symptoms after 3 and 5 days.

### Clinical presentation of encephalitis in COVID-19 disease

At onset, the most common symptoms were delirium/altered mental status (n=17, 68%), aphasia/dysarthria (n=6, 24%) and seizures (n=6, 24%).

During the disease course a total of 9 patients (36%) showed aphasia, 8 (32%) had dysarthria and 6 cases (24%) exhibited focal motor deficits. Seizures were detected in nine patients of whom four developed a focal status epilepticus. Only one patient developed a parkinsonian syndrome. Headache was reported at onset in 6 patients (24%) and by a total of 10 patients (40%) during the disease course. Two patients had a previous diagnosis of stroke, one of mental retardation (with no structural brain abnormalities) and one patient had a previous (2019) diagnosis of possible encephalitis and Behçet disease (negative testing for autoantibodies in serum and CSF and imaging not suggestive for neuro-Behçet). Twelve patients had a diagnosis of hypertension, 6 of dyslipidaemia and 4 of diabetes mellitus.

### EEG, CSF analysis and blood screening

EEG was abnormal in all cases, showing in most cases a generalized slowing especially localized on frontal derivation (64%), while focal epileptic alteration was observed in six cases (24%). CSF analysis was abnormal in 17/25 patients (68%): 15 (60%) patients showed hyperproteinorrachia (50 to 123 mg/dL) and 6 (24%) pleocytosis (5 to 55 cells). CSF screening for bacteria and virus infection was negative in all patients (Supplementary Table 1). Fourteen patients underwent CSF RT-PCR for SARS-CoV-2 and all cases were negative (in 5 patients repeated in a second CSF analyses).

Thyroid function and metabolic/infectious screening (supplementary table 1) was normal in all patients. Thirteen patients with suspected for autoimmune encephalitis resulted negative for antibodies to intracellular, synaptic, or surface antigens (Euroimmun kit, Luebeck, Germany - see Supplementary Table 1 for details).

### Brain MRI imaging

Brain MRI was performed with a median of 6 (IQR 4-9) days from the onset of neurological symptoms. As shown in tables 1 and 2 and figure 1, 13 patients (52%) had a normal MRI (E-MRI-), 5 (20%) patients showed specific MRI alterations (ADEM n=1, ANE n=2 and LE n=2), whereas 7 (28%) patients showed heterogeneous MRI alterations (E-MRI+) such as multiple subcortical T2-hyperintensities (n=4), focal cortical T2 and DWI hyperintensities (n=3) and leptomeningeal enhancement (n=1).

**Table 2.** Clinical and instrumental characteristics according to encephalitis subtypes.

|  | Encephalitis,<br>all (n=25) | ADEM /ANE<br>(n=3) | LE<br>(n=2) | E-MRI+<br>(n=7) | E-MRI-<br>(n=13) | p |
| --- | --- | --- | --- | --- | --- | --- |
| **Demographic characteristics** |  |  |  |  |  |  |
| Age, years | 65.9 ± 9.6 | 65.3 ± 9.5 | 72.0 ± 5.7 | 63.6 ± 10.4 | 66.5 ± 10.2 | 0.26 |
| Sex female, n (%) | 10 (40%) | 0 (0%) | 1 (50%) | 3 (42.9%) | 6 (46.2%) | 0.51 |
| **Symptoms at onset** |  |  |  |  |  |  |
| Delirium, n (%) | 17 (68%) | 3 (100%) | 0 (0%) | 4 (57.1%) | 10 (76.9%) | 0.45 |
| Seizures, n (%) | 6 (24%) | 0 (0%) | 1 (50%) | 2 (33.4%) | 3 (23.1%) | 0.11 |
| Aphasia, n (%) | 3 (12%) | 0 (0%) | 0 (0%) | 1 (14.3%) | 2 (15.4%) | 0.34 |
| Dysarthria, n (%) | 3 (12%) | 0 (0%) | 1 (50%) | 1 (14.3%) | 1 (7.7%) | 0.25 |
| **Symptoms during the disease course** |  |  |  |  |  |  |
| Delirium, n (%) | 19 (76%) | 3 (100%) | 1 (50%) | 5 (71.4%) | 9 (69.3%) | 0.45 |
| Seizures, n (%) | 9 (36%) | 0 (0%) | 1 (50%) | 5 (71.4%) | 3 (23.1%) | 0.17 |
| Aphasia, n (%) | 9 (36 %) | 2 (66.7%) | 1 (50%) | 2 (33.4%) | 4 (30.8%) | 0.15 |
| Dysarthria, n (%) | 8 (32%) | 1 (33.3%) | 1 (50%) | 2 (33.4%) | 4 (30.8%) | 0.57 |
| Focal symptoms, n (%) | 6 (24%) | 2 (66.7%) | 1 (50%) | 1 (14.3%) | 2 (15.4%) | 0.49 |
| Movement disorders, n (%) | 1 (4%) | 0 (0%) | 0 (0%) | 1 (14.3%) | 0 (0%) | 0.44 |
| **CSF** |  |  |  |  |  |  |
| Pathological findings, n (%) | 17 (68%) | 3 (100%) | 2 (100%) | 5 (71.4%) | 7 (53.8%) | 0.10 |
| Protein, mg/L | 60.1 ± 35.0 | 86.4 ± 33.7 | 56.5 ± 29.0 | 61.7 ± 37.1 | 53.7 ± 35.8 | 0.51 |
| Hyperproteinorrhachia, n (%) | 15 (60%) | 3 (100%) | 1 (50%) | 4 (57.1%) | 7 (53.8%) | 0.28 |
| Cells count/uL, mean (range) | 6.4 ± 11.8 | 2.3 ± 2.5 | 8.0 ± 9.9 | 11.6 ± 19.9 | 4.4 ± 6.7 | 0.17 |
| Pleocytosis, n (%) | 9 (36%) | 1 (33%) | 1 (50%) | 3 (42.9%) | 4 (30.8%) | 0.50 |
| Glucose, mg/dL | 76.2 ± 25.0 | 72.3 ± 17.3 | 110.5 ± 71.4 | 81.6 ± 29.5 | 69.0 ± 9.0 | 0.21 |
| **Outcome** |  |  |  |  |  |  |
| Length of hospitalization (days) | 16.8 ± 8.15 | 17.3 ± 2.5 | 19.0 ± 4.2 | 19.8 ± 11.1 | 14.7 ± 7.61 | 0.15 |
| Median mRS pre-admission | 0.6 ± 0.8 | 0.7 ± 1.1 | 0.0 | 0.9 ± 0.9 | 0.5 ± 0.8 | 0.77 |
| Median mRS at discharge | 2.4 ± 1.9 | 4.0 ± 1.7 | 1.0 ± 1.4 | 4.3 ± 2.1 | 1.8 ± 1.7 | **0.03** |
| Death, n (%) | 4 (16%) | 0 (0%) | 0 (0%) | 3 (42.8%) | 1 (7.70%) | 0.17 |

**Abbreviations:** ADEM, acute disseminated encephalomyelitis; ANE, acute necrotizing encephalopathy; CSF, cerebrospinal fluid; E-MRI-, unknown origin encephalitis with negative MRI; E-MRI+, unknown origin encephalitis with MRI alterations; LE, limbic encephalitis; mRS, modified Ranking Scale. p value indicated Fisher exact test and ANOVA test for dichotomic and continuous variables, respectively.

**Figure 1.**
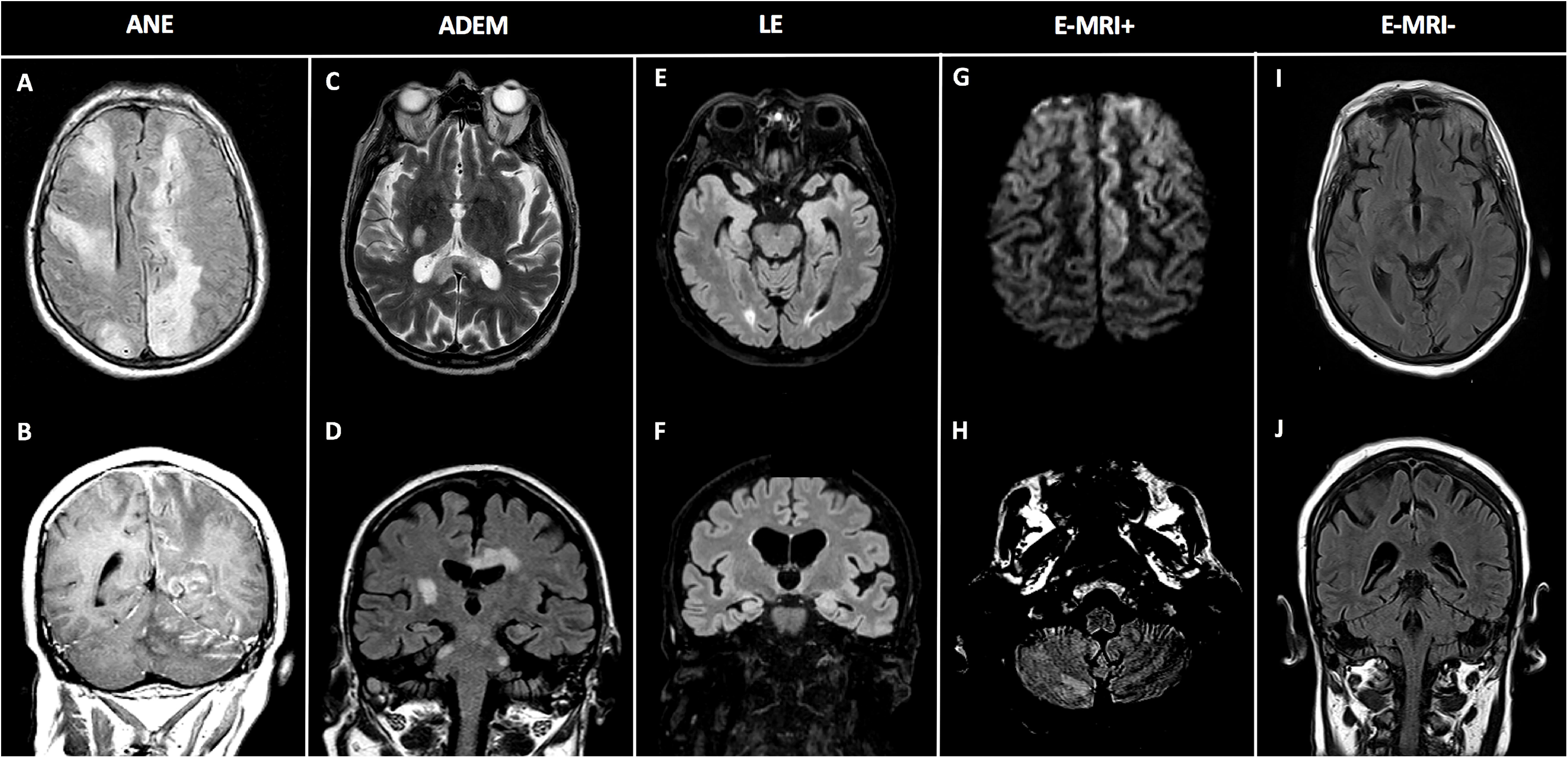
Brain Magnetic Resonance Imaging findings according to encephalitis phenotypes. Panel A-B: A case of acute necrotizing encephalitis (ANE) characterized by FLAIR diffuse bilateral hyperintensities and (A), with linear Gadolinium-enhancement on coronal T1 (B). Panel C-D: A case of ADEM with T2 and FLAIR hyperintensities (involving corpus callosum, bilateral cerebellar peduncles and right thalamus) on axial (C) and coronal plane (D). Panel E-F: A case of limbic encephalitis (LE) characterized by increased T2-FLAIR signal within bilateral mesial temporal lobes and (E) and coronal (F) planes. Panel G-H: A case of unspecific alterations defining the group of E-MRI+: DWI hyperintensities on frontal superior and medium gyrus and FLAIR hyperintensity on right cerebellar tonsil. Panel I-J A case of E-MRI-without pathological findings on FLAIR images. **Abbreviations:** ADEM, acute disseminated encephalomyelitis; ANE, acute necrotizing encephalitis; E-MRI-, encephalitis with negative MRI; E-MRI+, encephalitis with MRI alterations; LE, Limbic encephalitis.

### Clinical presentation according to encephalitis subtypes

All ADEM and LE cases occurred after the onset of COVID-19 symptoms with a delayed neurological onset compared with the remaining encephalitis cases. COVID-19 respiratory involvement was significantly more severe in ADEM/ANE and LE compared to other encephalitis (Table 1).

Blood and CSF analyses did not differ between encephalitis’ subgroups but higher LDH levels were found in ADEM and LE cases (p=0.001, supplementary Table 2). Clinical presentation at onset and during the hospitalisation were similar between groups, but a higher frequency of seizures was observed in E-MRI+ cases (Table 2).

Spontaneous recovery was observed in 6 patients. A positive response to high-dose methylprednisolone was observed in one ADEM, in one LE and in 3 E-MRI-patients, whereas no response was observed in ANE and all four E-MRI+ cases. Immunoglobulins (1 g/Kg for five days) were administered to one ANE, two E-MRI+ and one E-MRI-without significant benefits. The final clinical outcome was worse in ADEM/ANE and E-MRI+ cases compared to LE and E-MRI-encephalitis (p=0.03, Figure 2). Four cases deceased, namely 3 E-MRI+ and one E-MRI-.

**Figure 2.**
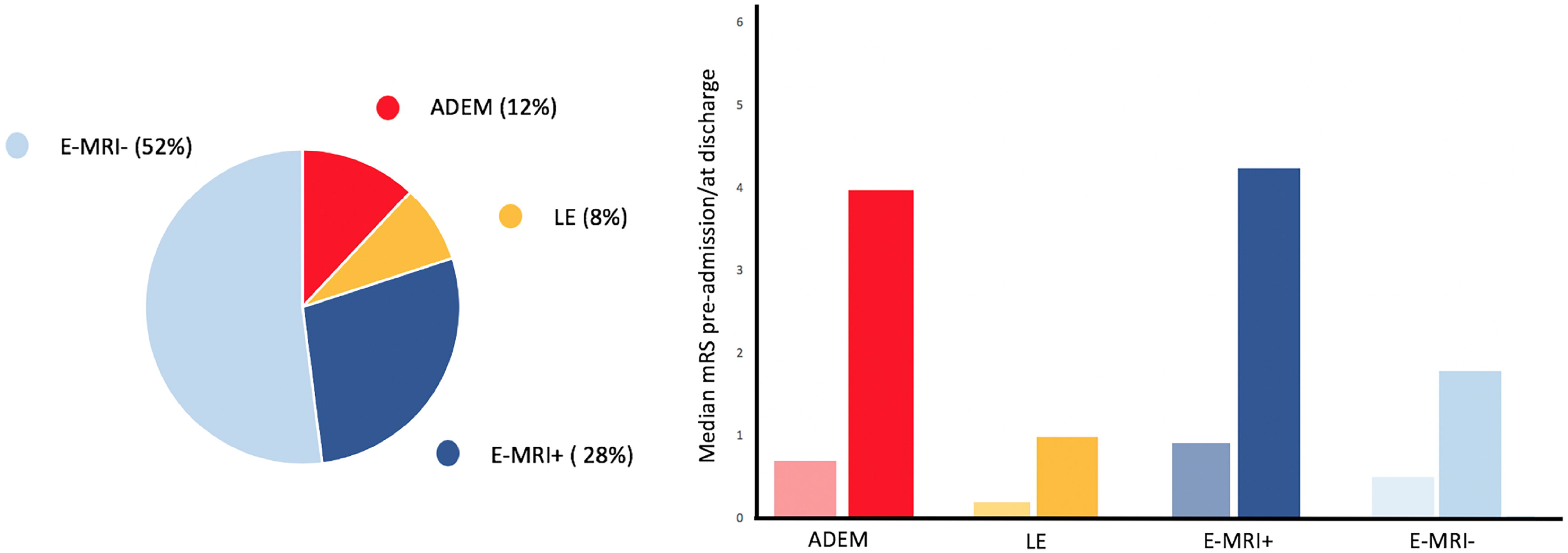
Outcome measures according to the encephalitis subtypes. The histograms showed premorbid mRS (light colors) and final mRS (enhanced colors). **Abbreviations:** ADEM, acute disseminated encephalomyelitis; E; E-MRI-, encephalitis with negative MRI; E-MRI+, encephalitis with MRI alterations; LE, limbic encephalitis; mRS modified Ranking scale.

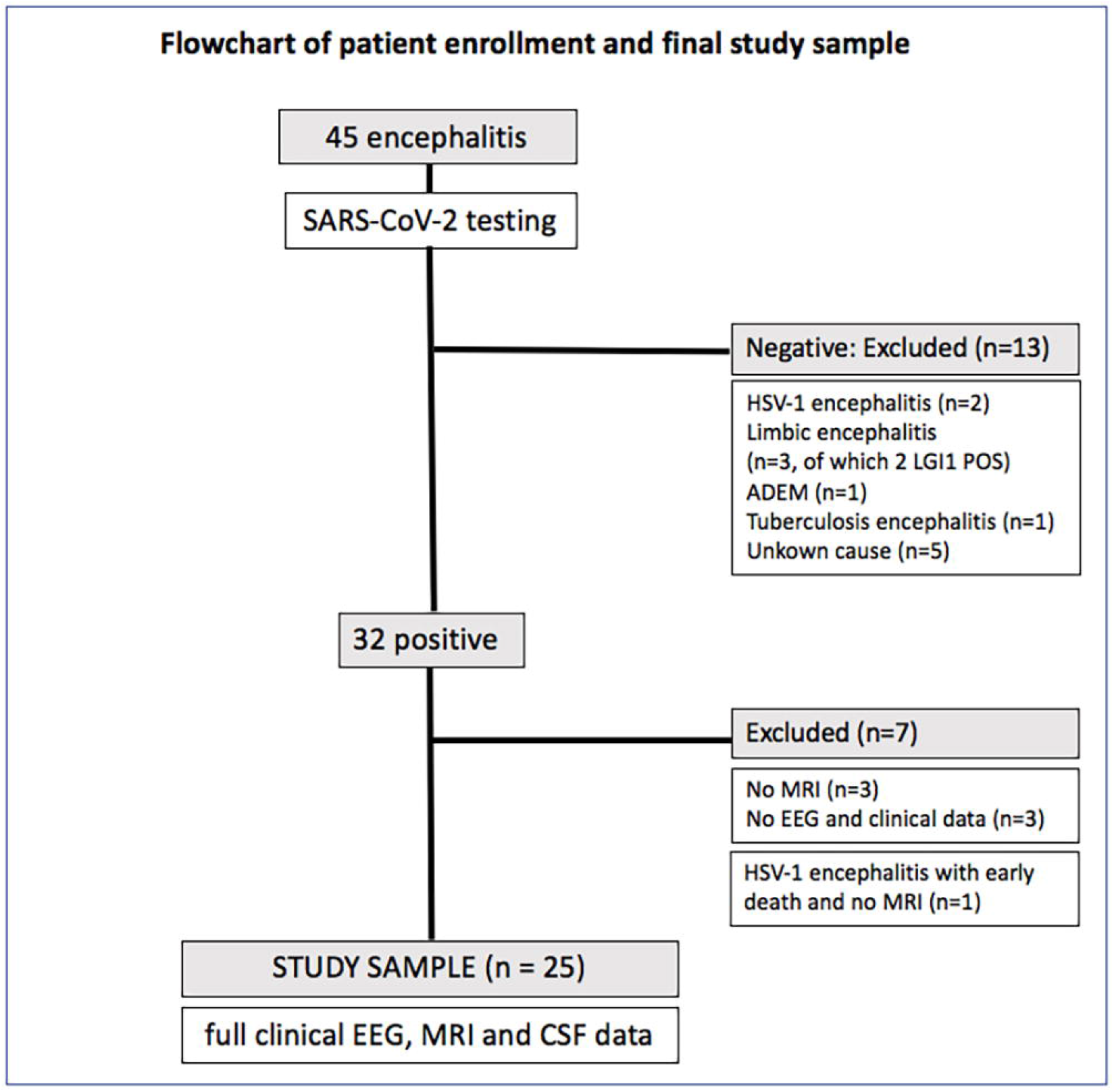

## Discussion

In this multicentre study, SARS-CoV-2 infection was associated with a wide clinical spectrum of encephalitis, characterised by heterogeneous clinical presentation and outcomes, thus underlying divergent pathophysiological mechanisms.

The cohort included 25 cases of encephalitis diagnosed according to standard guidelines^13^ with confirmed SARS-CoV-2 respiratory infection and full CSF, EEG, MRI available data, recruited in a network involving 13 centres in Lombardy and Veneto, Italy.

The large majority of patients exhibited moderate respiratory COVID-19 disease, with prominent neurological disturbances including delirium and aphasia/dysarthria as most common symptoms at onset. Seizures were reported in about a third of patients during disease course with few cases developing status epilepticus, in line with presentation of encephalitis reported in large multicentre studies.^20,21^ The CSF analyses showed mild pleocytosis and/or hyperproteinorrachia in two-third of cases though viral and bacterial screening was negative in all patients. Nineteen CSF specimens resulted negative for SARS-CoV-2 virus using RT-PCR technique.

To date, only two cases of encephalitis reported the presence of SARS-COV-2 in CSF, despite negative peripheral and respiratory findings^24^ (supplementary Table 3), and viral potential diffusivity was also demonstrated for similar viral agents such as SARS-CoV.^22,23^

It should be remarked that the absence of virus in CSF could not definitely exclude a direct viral invasion, as demonstrated for other infectious diseases such as West Nile virus or enterovirus infections.^13^ However, the clinical courses and CSF alterations in this series of patients argue against a direct CNS damage and conversely claim for neuroinflammatory and, in rarer cases, autoimmune responses as major players in COVID-19 encephalitis.

This hypothesis was further supported by the cases-classification based on MRI findings. In fact, the majority of patients exhibited normal imaging or heterogeneous T2-hyperintensities in cortical/subcortical areas. Further, we identified three cases of ADEM and two cases of LE, thus supporting an immune-mediated-mechanism.

Acute disseminated encephalomyelitis has been already reported after several viral infections, including SARS-COV, MERS-COV^25^ and SARS-CoV-2.^26^ In our study, two patients presented with the acute necrotising encephalitis (ANE), a severe post-infectious paediatric complication - reported in a single SARS-CoV-2 patient by Poyadji and coauthors so far.^25^ Two additionally cases showed typical MRI findings consistent with limbic encephalitis, known to be associated in the majority of cases with autoimmune or paraneoplastic diseases,^13^ despite we did not find any known specific antibodies in biofluids.^16^ The onset of ADEM/LE cases was delayed after the onset of COVID-19 symptoms, that were more severe in terms of respiratory function compared to other encephalitis. Immuno-modulatory treatment indeed showed high efficacy in the typical ADEM and LE cases, while ANE presented more severe final outcomes-mostly related to known haemorrhagic complications.^18^

The other twenty cases of encephalitis occurred almost concomitantly to SARS-CoV-2 respiratory infections, in the majority of cases few days after the onset of COVID-19 symptoms. Two-thirds exhibited negative MRI with CSF abnormalities, in line with cases described for SARS-COV2,^4,5,7,11^ but also for SARS-CoV^26^ and MERS-CoV infections.^11,22,23^ We also observed a subset of patients with heterogeneous MRI alterations. These patients showed similar clinical onset and presentations to other phenotypes though they displayed higher prevalence of epileptic seizures. The close time-relationship with SARS-CoV2 infection in these cases argues for a CNS involvement via inflammatory-mediated mechanisms. Recent reports claimed for a cytokine-mediated neuroinflammation in COVID-19 encephalitis,^27^ as an increased IL-8/TNF-alpha in CSF was observed in one patient with negative MRI findings^6^ and two cases with temporal lobe/splenium abnormalities.^28^ This cytokine-mediated neuroinflammation would fit with the response to immunomodulation treatment observed in some cases and the spontaneous recovery of milder presentations, especially those with negative MRI findings. This is in line with different degree of neurotoxicity of cytokine-induced syndrome observed after immune effector cell-associated treatments.^29^ Better response to treatment and final outcome was indeed reported in patients with negative neuroimaging compared to patients with MRI alterations, who in turn exhibited a higher mortality rate.

The wide spectrum of clinical presentation of encephalitis in patients with SARS-CoV2 respiratory infection is similar with phenotypes already associated with influenza and other coronavirus.^11,22,23,26,30^ The neurological complications of influenza, however, have an estimated incidence between 0.21 and 12 cases per million and affects especially children.^30^ Based on our data on about 3 months of observation, we estimated an incidence of encephalitis in COVID-19 of at least 58/100.000 cases. These findings are in agreement with still unpublished reports on COVID-19 neurological presentation in UK^30^ and the trends of CNS complications in SARS and MERS recently proposed by Ellul and coauthors,^11^ who estimated an incidence between 37 and 224 cases per 100.000 symptomatic patients.

Overall, the proportion of patients with neurological disease is likely to remain small compared with respiratory symptoms in COVID-19 disease. Nevertheless, as the pandemic continues, the overall number of encephalitis will probably increase both in the acute and in the post-acute phase. This should encourage larger consortia and studies evaluating encephalitis in COVID-19 disease in order to develop specific diagnostic algorithm and possibly different management strategies according to single-case presentations.

We acknowledge that this study entails several limitations. First, due to the absence of autopsy or SARS-CoV-2 evidence in CSF, our cases can be classified as “probably associated” with SARS-CoV-2 infection^11^ and further studies on CSF using recently-developed intrathecal antibodies are urgently needed. Second, the study might have underestimated the total number of cases, especially severe cases who could not undergo MRI, EEG and CSF assessment, likely due to the prominent respiratory insufficiency with early intubation and/or death. Third, the extensive panel of paraneoplastic/autoimmune encephalitis was not performed in about a third of patients exhibiting transitory symptomatology albeit MRI/CSF was not suggestive for immune disorders. Despite these limitations, this is the first multicentre study including the highest number of SARS-CoV-2 cases fulfilling criteria for encephalitis.^13^

## Conclusions

SARS-CoV-2 infection is associated with a wide spectrum of encephalitis. The majority of cases were concomitant to SARS-CoV-2 infection with findings arguing for a cytokine-mediated CNS-specific involvement. The few cases of ADEM/ LE syndromes, characterized by a delayed in onset and more severe respiratory distress, were likely linked to immuno-mediated mechanisms.

Further studies focusing on biomarkers are needed in order to disentangle the pathophysiological mechanisms underlying the wide spectrum of CNS involvement in COVID-19 disease and to find the best management strategies for this worldwide growing condition.

## Data Availability

The data are available from the author upon reasonable request

## Author contribution

Conception and design of the study: AP and AP. Acquisition and analysis of data: AP, SM, IV, EZ, EM, VG, ML, FC, NR, MS, MG, MSC, MT, UB, SM, SF, AC, FF, AI, BR, AB, EP, ML, EF, FC, ML, RG, FC, GZ, AP and AP.

Drafting the manuscript and figures: AP, SM and AP.

## Potential Conflict of interest

Andrea Pilotto is consultant and served on the scientific advisory board of Z-cube (Technology Division of Zambon Pharma), received speaker honoraria from Biomarin and Zambon Pharmaceuticals.

SM, IV, EZ, EM, VG, ML, FC, NR, MS, MG, MSC, MT, SM, SF, AC, FF, AI, BR, AB, EP, ML, EF, FC, ML, RG, FC, GZ, AP reported no disclosures.

Alessandro Padovani is consultant and served on the scientific advisory board of GE Healthcare, Eli-Lilly and Actelion Ltd Pharmaceuticals, received speaker honoraria from Nutricia, PIAM, Lansgstone Technology, GE Healthcare, Lilly, UCB Pharma and Chiesi Pharmaceuticals.

## #The ENCOVID study group includes

Andrea Pilotto^1,2^ MD, Stefano Masciocchi^1^ MD, Irene Volonghi^1^ MD, Elisabetta del Zotto^3^ MD, Massimo Crabbio^3^ MD, Eugenio Magni^3^ MD, Valeria De Giuli^4^ MD, Francesca Caprioli^4^ MD, Nicola Rifino^5^ MD, Maria Sessa^5^ MD, Michele Gennuso^6^ MD, Maria Sofia Cotelli^7^ MD, Marinella Turla^7^ MD, Ubaldo Balducci^8^ MD, Sara Mariotto^9^ MD, Sergio Ferrari MD^9^ MD, Alfonso Ciccone^10^ MD, Fabrizio Fiacco^11^ MD,, Massimiliano Guindani^12^ MD, Alberto Imarisio^1^ MD, Barbara Risi^1^ MD, Alberto Benussi^1^ MD, Loris Poli^1^ MD, Stefano Gipponi^1^ MD, Massimiliano Filosto^1^ MD, Enrico Premi^13^ MD, Massimo Gamba^13^ MD, Salvatore Caratozzolo^1^ MD, Viviana Cristillo^1^ MD, Ilenia Libri^1^ MD, Francesca Schiano di Cola^1^ MD, Stefano Cotti Piccinelli^1^ MD, Matteo Cortinovis^1^ MD, Andrea Scalvini^1^ MD Enrico Baldelli^1^ MD, Martina Locatelli^1^ MD, Matteo Benini^14^ MD, Stefano Gazzina^15^ MD, Erika Chiari^16^ MD, Silvia Odolini^16^ MD, Emanuele Focà^16^ MD, Francesca Caccuri^17^ MD, Arnaldo Caruso^17^ MD, Matilde Leonardi^18^ MD, Claudia Ambrosi^19^ MD, Lorenzo Pinelli^19^ MD, Roberto Gasparotti^19^ MD, Simonetta Gerevini^20^, Elisa Francesca Maria Ciceri^21^ MD, Francesco Castelli^16^ MD, Gianluigi Zanusso^7^ MD, Bruno Ferraro^22^ MD, Giorgio Dalla Volta^23^ MD, Alessandro Pezzini^1^ MD, Alessandro Padovani^1^ MD PhD on behalf of ENCOVID research group

^1^Neurology Unit, Department of Clinical and Experimental Sciences, University of Brescia, Italy

^*2*^ Parkinson’s Disease Rehabilitation Centre, FERB ONLUS – S. Isidoro Hospital, Trescore Balneario (BG), Italy

^3^ Neurology Unit, Poliambulanza Hospital, Brescia, Italy

^4^ Neurology Unit, Istituti Ospedalieri, ASST Cremona, Cremona, Italy

^5^ Department of Neurology, Papa Giovanni XXIII Hospital, ASST Papa Giovanni XXII, Bergamo, Italy

^6^ Neurology Unit, Crema Hospital, Crema, Italy

^7^ Neurology Unit, ASST Valcamonica, Esine, Brescia, Italy

^8^ Neurology Unit, ASST Chiari, Chiari, Italy

^9^ Neurology Unit, Department of Neurosciences, Biomedicine, and Movement Sciences, University of Verona, Verona, Italy

^10^ Department of Neurology and Stroke Unit, Carlo Poma Hospital, ASST Mantova, Mantova, Italy

^11^ Neurology Unit, ASST Bergamo Est, Seriate, Italy ^12^Neurology Unit, Sant’Anna Hospital, Brescia, Italy ^13^ Stroke Unit, ASST Spedali Civili di Brescia, Italy

^14^ Neurology Unit, School of Medicine, University of Bologna, Bologna, Italy

^15^ Neurophysiology Unit, ASST Spedali Civili of Brescia, Italy

^16^ University Division of Infectious and Tropical Diseases, University of Brescia and ASST Spedali Civili Hospital, Brescia, Italy

^17^ Microbiology Unit, Department of Molecular and Translational Medicine, University of Brescia and ASST Spedali Civili Hospital, Brescia, Italy

^18^ Neurology, Public Health, Disability Unit - IRCCS Neurology Institute Besta, Milan, Italy

^19^Neuroradiology Unit, Department of Molecular and Translational Medicine, University of Brescia and ASST Spedali Civili Hospital, Brescia, Italy

^20^ Neuroradiology Unit, Papa Giovanni XXIII Hospital, ASST Papa Giovanni XXII, Bergamo, Italy

^21^ Neuroradiology Unit, University of Verona, Verona

^22^ Neurology Unit, ASST Bergamo Ovest, Treviglio, Italy

^23^ Neurology Department, Città di Brescia Institute, Brescia, Italy

## Notes

### Competing Interest Statement

The authors have declared no competing interest.

### Funding Statement

The study was not funded.

### Author Declarations

Neurology Unit, Department of Clinical and Experimental Sciences University of Brescia

